# Rare diseases load through the study of a regional population

**DOI:** 10.1101/2024.10.29.24316346

**Authors:** Élisa Michel, Claudia Moreau, Laurence Gagnon, Josianne Leblanc, Jessica Tardif, Lysanne Girard, Jean Mathieu, Cynthia Gagnon, Mathieu Desmeules, Jean-Denis Brisson, Luigi Bouchard, Simon L. Girard

**Affiliations:** Département des sciences fondamentales, Université du Québec à Chicoutimi, Saguenay, Québec, Canada; Centre Intersectoriel en Santé Durable (CISD), Université du Québec à Chicoutimi, Saguenay, Québec, Canada; Département clinique de médecine de laboratoire du Centre intégré universitaire de santé et services sociaux (CIUSSS) du Saguenay–Lac-St-Jean, Saguenay, Québec, Canada; Département de biochimie et de génomique fonctionnelle, Faculté de médecine et des sciences de la santé, Université de Sherbrooke, Saguenay, Québec, Canada; Groupe de recherche interdisciplinaire sur les maladies neuromusculaires (GRIMN), CIUSSS du Saguenay–Lac-Saint-Jean, Saguenay, Québec, Canada; Faculté de médecine et des sciences de la santé, Université de Sherbrooke, Saguenay, Québec, Canada; Centre de recherche et d’innovation du CIUSSS du Saguenay–Lac-St-Jean, Saguenay, Québec, Canada; Clinique de pédiatrie du Saguenay, Saguenay, Québec, Canada; Clinique des maladies neuromusculaires (CMNM), CIUSSS du Saguenay–Lac-St-Jean, Saguenay, Québec, Canada; Projet BALSAC, Université du Québec à Chicoutimi, Saguenay, Québec, Canada; Centre de recherche CERVO, Université Laval, Québec, Québec, Canada

## Abstract

Rare genetic diseases impact many people worldwide and are challenging to diagnose. In this study, we introduce a novel regional population cohort approach to identify pathogenic variants that occur more frequently within specific populations and are of clinical interest. We utilized a cohort from Quebec, including the Saguenay–Lac-Saint-Jean region, which is known for its founder effect and higher frequency of certain pathogenic variants. By analyzing both the frequency of these variants and their origin through shared identical-by-descent segments, we validated 38 variants previously reported as being more common due to the founder effect. Additionally, we identified 42 unreported founder variants in Quebec or the Saguenay–Lac-Saint-Jean, some with carrier rates estimates as high as 1/22. We also observed a greater deleterious mutational load for the studied variants in individuals from the Saguenay–Lac-Saint-Jean compared to other urban Quebec regions. These findings were brought to the clinic where 12 pathogenic variants were detected in patients, including 3 that are responsible for very severe diseases and could be considered for inclusion in a carrier test for the Saguenay–Lac-Saint-Jean population. This study highlights the potential underestimation of rare disease prevalence and presents a population-based approach that could aid clinicians in their diagnostic efforts and patients’ management.

## Introduction

Rare diseases are thought to collectively affect as much as 10% of the population^1^. There are more than 10,000 rare diseases described in Orphanet^2^ and most of them are of genetic origin. Diagnosis remains a significant challenge for patients living with a rare disease. Despite the growing accessibility of genome sequencing technologies in precision medicine efforts for rare diseases diagnosis^3^, these patients often experience prolonged diagnostic odyssey due to insufficient knowledge about their specific condition and the diversity of symptoms observed for a given disease. It becomes increasingly important to improve the diagnostic yield of rare diseases and to shorten the diagnostic odyssey of patients^4^. Understanding population health disparities is an essential component of equitable precision health efforts.

In certain populations, the prevalence of some rare diseases may increase due to demographic events such as founder effects. It is the case in Quebec, a province in eastern Canada, predominantly settled by people of French origin starting in the early 1600s^5^. The initial European founder effect was followed by subsequent regional founder effects, notably the well-characterized one observed in Charlevoix and Saguenay–Lac-Saint-Jean (SLSJ) regions^6^. Consequently, many rare diseases are more frequent in SLSJ than elsewhere in the world^7– 10^. In SLSJ, most people are aware of the higher risk of transmission of some rare diseases and a carrier test is offered to the populations of Charlevoix, SLSJ and Haute-Côte-Nord for 4 of these diseases^8,11^. Nevertheless, numerous rare diseases still lack a known genetic etiology and diverse manifestations of diseases across patients further complicate clinical diagnosis. Traditionally, founder effects have been analyzed using a bottom-up approach, starting with the phenotypes of patients observed in clinical settings and linking them to genes that are specific to each individual. Often, medical geneticists and the healthcare system would gain valuable insights from obtaining a comprehensive overview of variants that are more frequent in the population and potentially associated with rare diseases. This study focuses on addressing this need.

More specifically, we aimed to describe potentially pathogenic variants that have an increased frequency in SLSJ due either to the founder effect or simply due to many introductions in the population. We conducted a comprehensive screening to identify pathogenic variants with higher frequency in SLSJ. Since the SLSJ population has been extensively studied over the past 40 years, we expected to identify many previously reported variants, thereby validating our findings. In fact, we successfully replicated and confirmed the majority of known founder variants in the SLSJ population and systematically documented their carrier rates. However, we also identified several variants that may be causal of rare diseases and were not previously documented in SLSJ. As the SLSJ healthcare system features a single entry point for all residents, it simplifies the process of locating patients with newly identified pathogenic variants. A thorough investigation of these newly discovered variants revealed clear diagnoses in the phenotypes of several patients.

Furthermore we report for the first time the global load of rare variants in a single population and assess how the founder effect was pivotal in increasing that load. In the context of rare diseases, a large number of populations remain poorly characterized and we believe that our study highlights the need for regional genetic programs to better understand and diagnose the variety of rare diseases affecting one population.

## Results

We detected 1,302 potentially pathogenic rare variants (Supplementary Table 1, see methods) in the whole genome sequencing (WGS) of 1,852 individuals within the Quebec province (QcP) that reached at least 10% relative frequency difference (RFD≥10%, see methods) compared to gnomAD non-Finnish Europeans. To improve carrier rate estimates, we imputed 29,353 individuals from CARTaGENE^12^ using the WGS as reference (see methods) and looked at these 1,302 variants. Whenever a variant was present in the imputed data, we used the imputed variant frequency, otherwise, we relied on the WGS variant frequency. The clustering on a Uniform Manifold Approximation and Projection (UMAP) performed on the imputed data identified 3,589 and 21,472 individuals who were genetically related to the SLSJ and the urban Quebec areas respectively (UQc, see methods). Noticeably, 540 (42%) variants with an RFD≥10% are absent from the SLSJ region and 17 (1.3%) only are absent from the UQc, although many variants are more frequent in the SLSJ region (Fig.1). Accordingly, we observed a lower proportion of individuals from the SLSJ that do not carry any potentially pathogenic variant (chi^2^ p-value < 2e^−16^) while a higher proportion carry 2 or more (chi^2^ p-value < 2e^−16^) (Fig.2).

**Fig. 1:**
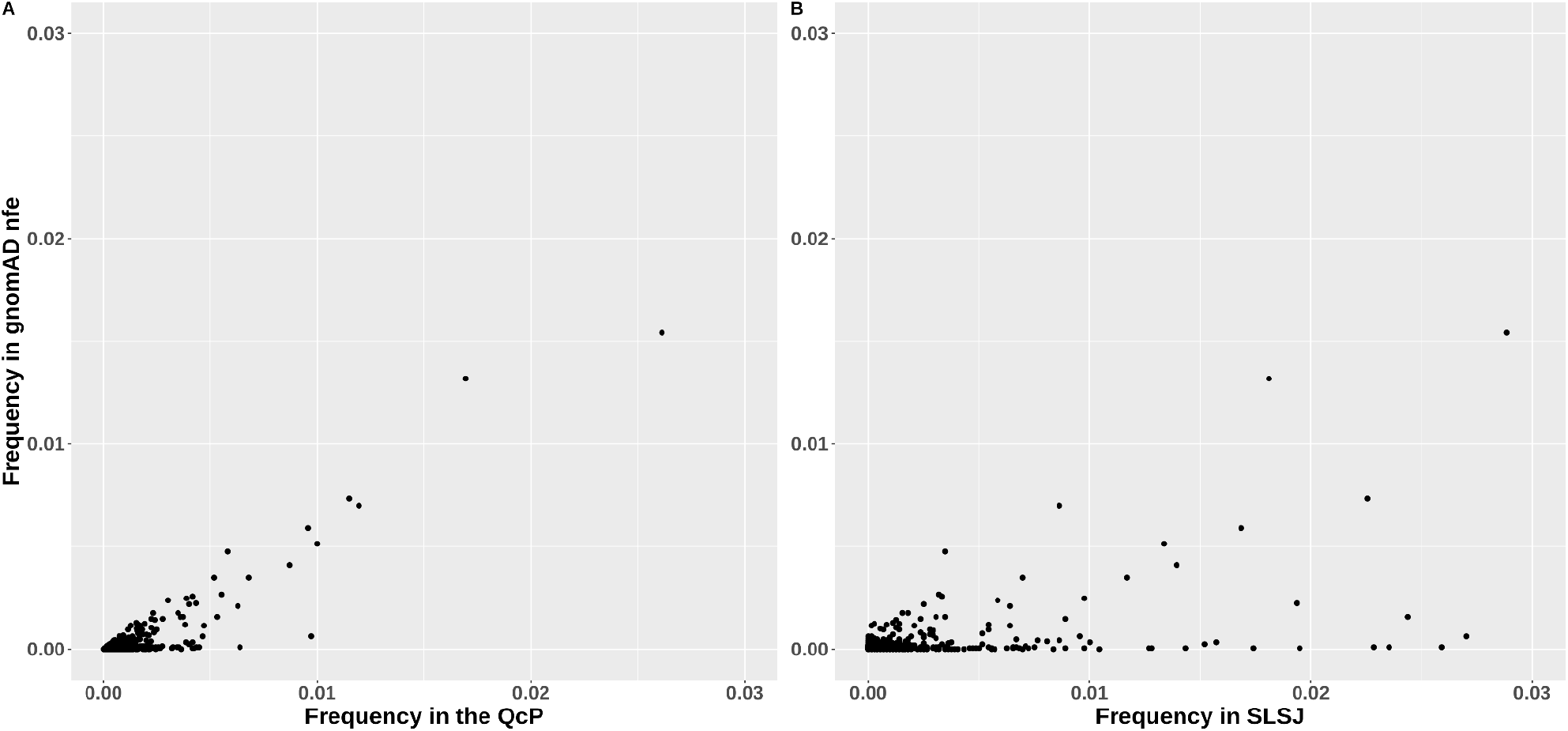
Frequencies of rare variants with RFD≥10% compared to gnomAD in A) QcP and B) SLSJ.

**Fig. 2:**
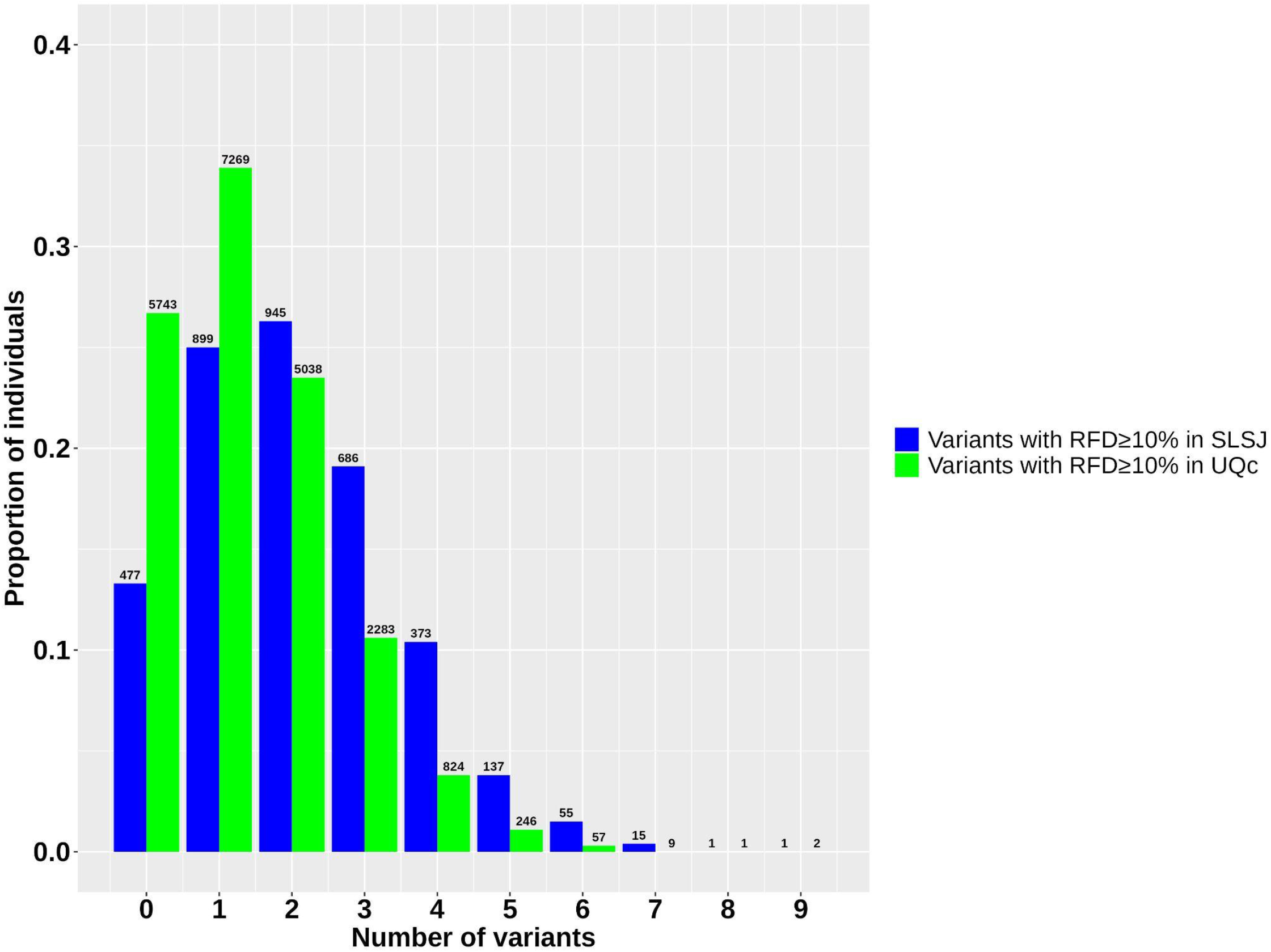
Proportion of individuals carrying variants with RFD≥10%.

### Previously reported and newly discovered variants

Previous literature reviews focussed on the Charlevoix-SLSJ founder effect identified 72 variants^7–10^. Among them, 42 were present in our data (Table 1 and Supplementary Table 1). Supplementary Table 2 provides details on the 30 previously reported variants that were either absent in our data or had an RFD<10%, as well as information on some variants that were not considered in our analysis. Noticeably, there is a great correlation between the carrier rates (CR) previously reported and the ones calculated herein (Supplementary Fig.1). Moreover, some carrier rates were reassessed in the CIUSSS laboratory using a subset of 1,000 randomly selected samples with the appropriate consent, and the newly calculated rates fall within the range of those reported in this study (Supplementary Table 3).

**Table 1:**
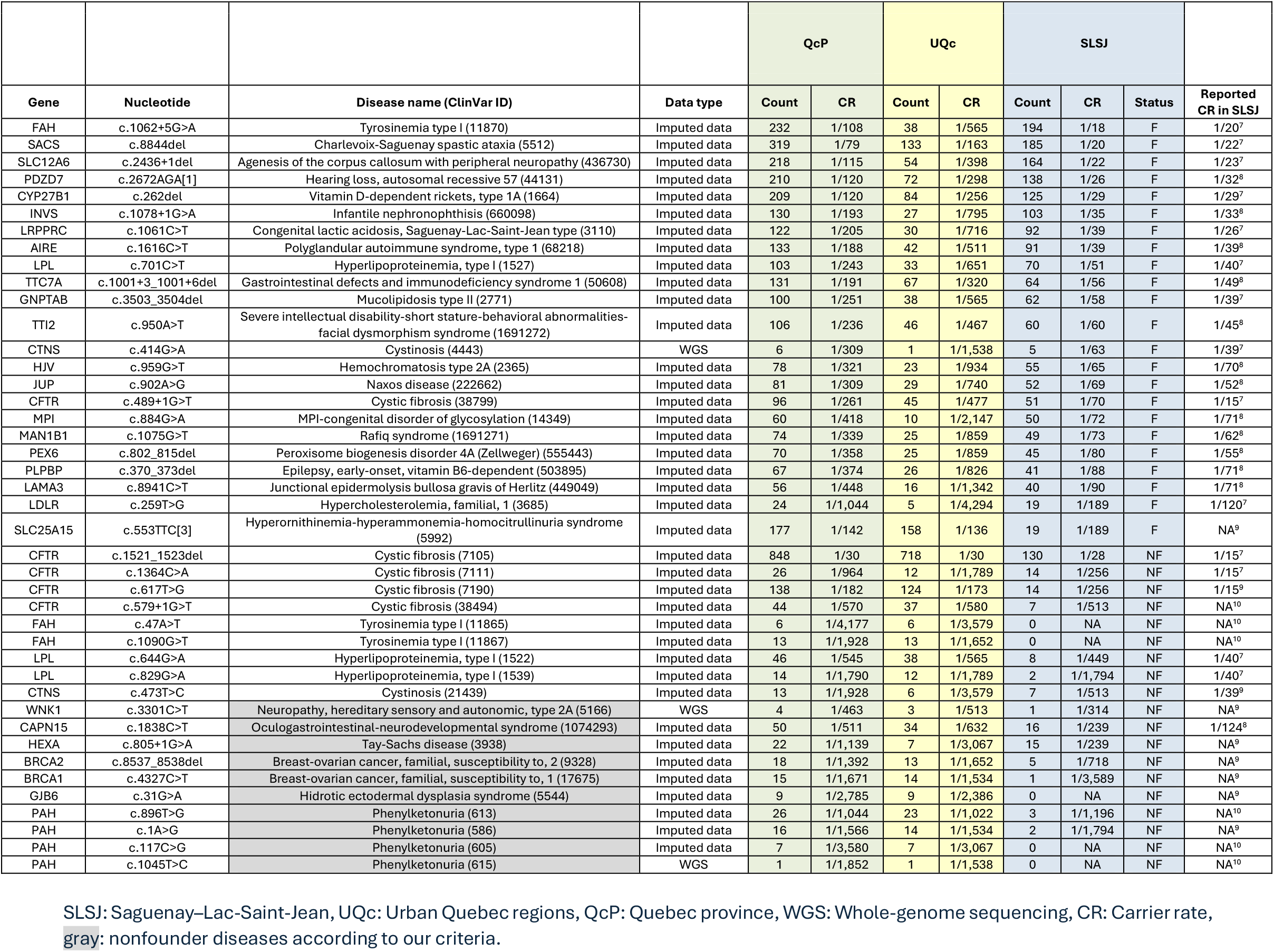
Variants previously reported.

In the present study, to be classified as founder, the variant must be present with a carrier rate of at least 1/200 and the proportion of pairs of carriers sharing segments identical-by-descent (IBD) around the variant should be at least 0.5 (see methods). Among the 1,302 rare variants with RFD≥10%, 80 variants met these criteria and are considered as founders either in the QcP, UQc or SLSJ. We reviewed the literature on these 80 variants, examining not only the reviews focusing on the Charlevoix-SLSJ founder effect^7–10^, but also case reports within the QcP population. While these reports did not primarily emphasize the founder effect, we still classified the variants therein as previously reported^13–27^. 38 of the founder variants were already documented whereas 42 were not reported in the Quebec population (Table 2 and Supplementary Table 1).

### Founder variants’ regional carrier rates and individuals’ mutation load

We then compared the carrier rates between the SLSJ and UQc (Fig.3). Most of the already reported founder variants are at higher CR than the newly identified ones, but some of the latters are as high as 1/22 in the SLSJ (Table 2). Carrier rates are generally higher in the SLSJ compared to the UQc. Specifically, the count of variants with carrier rates higher than 1/200 is 8 times higher in SLSJ than in the UQc (3 times higher when considering all variants with an RFD≥10% regardless of whether they are founder variants) (Fig.4). Consequently, the number of individuals who carry at least one potentially pathogenic founder variant is higher in the SLSJ than in the UQc (chi^2^ p-value < 2e^−16^) (Fig.5). In fact, for the variants already reported in the literature, 50% of the SLSJ and only 11% of the UQc individuals carry at least one variant. Notably, when the newly identified variants are added, these percentages reach 66% and 18%, respectively (chi^2^ p-value < 2e^−16^).

**Table 2:**
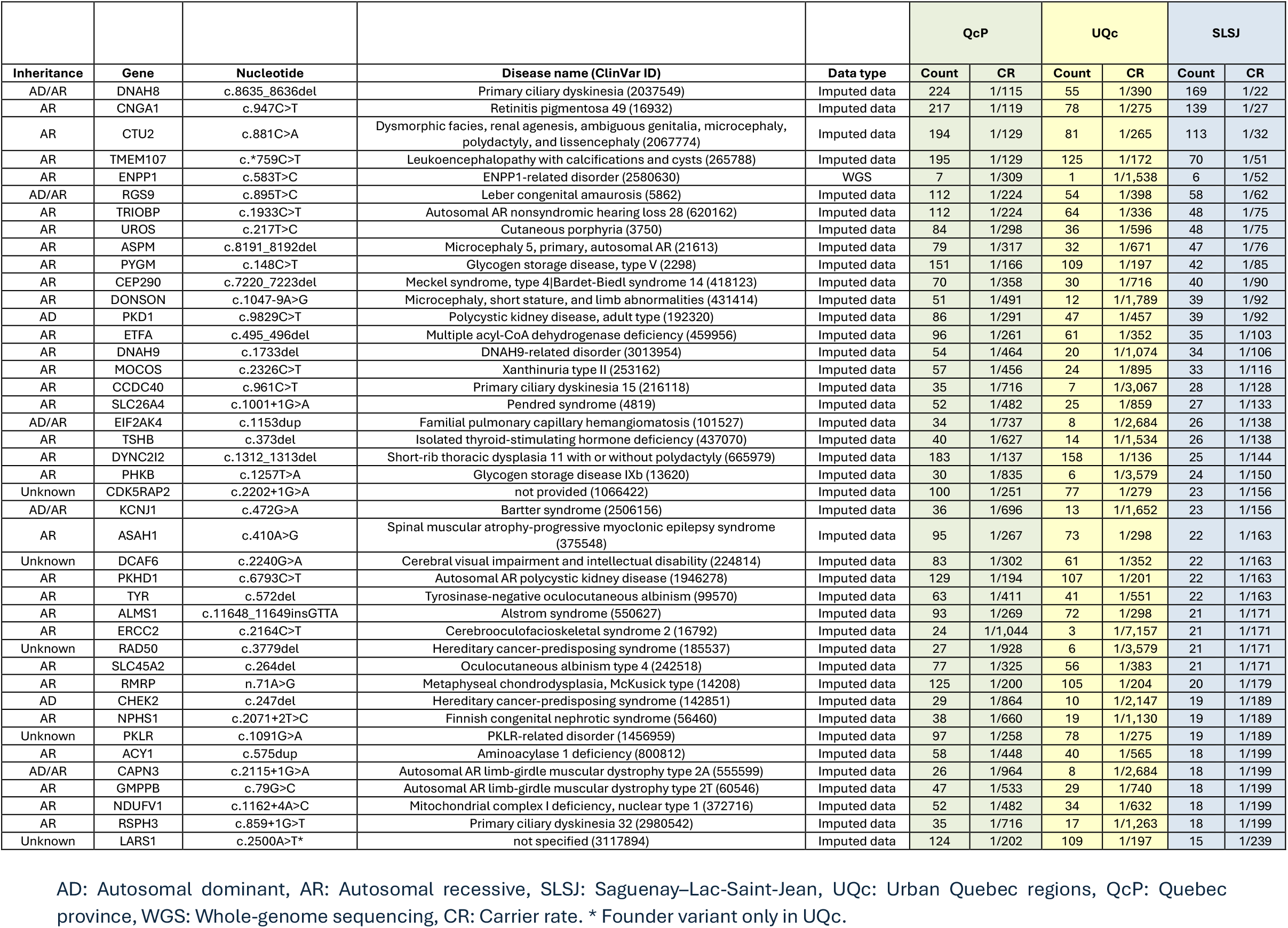
Novel founder variants found in this study.

**Fig. 3:**
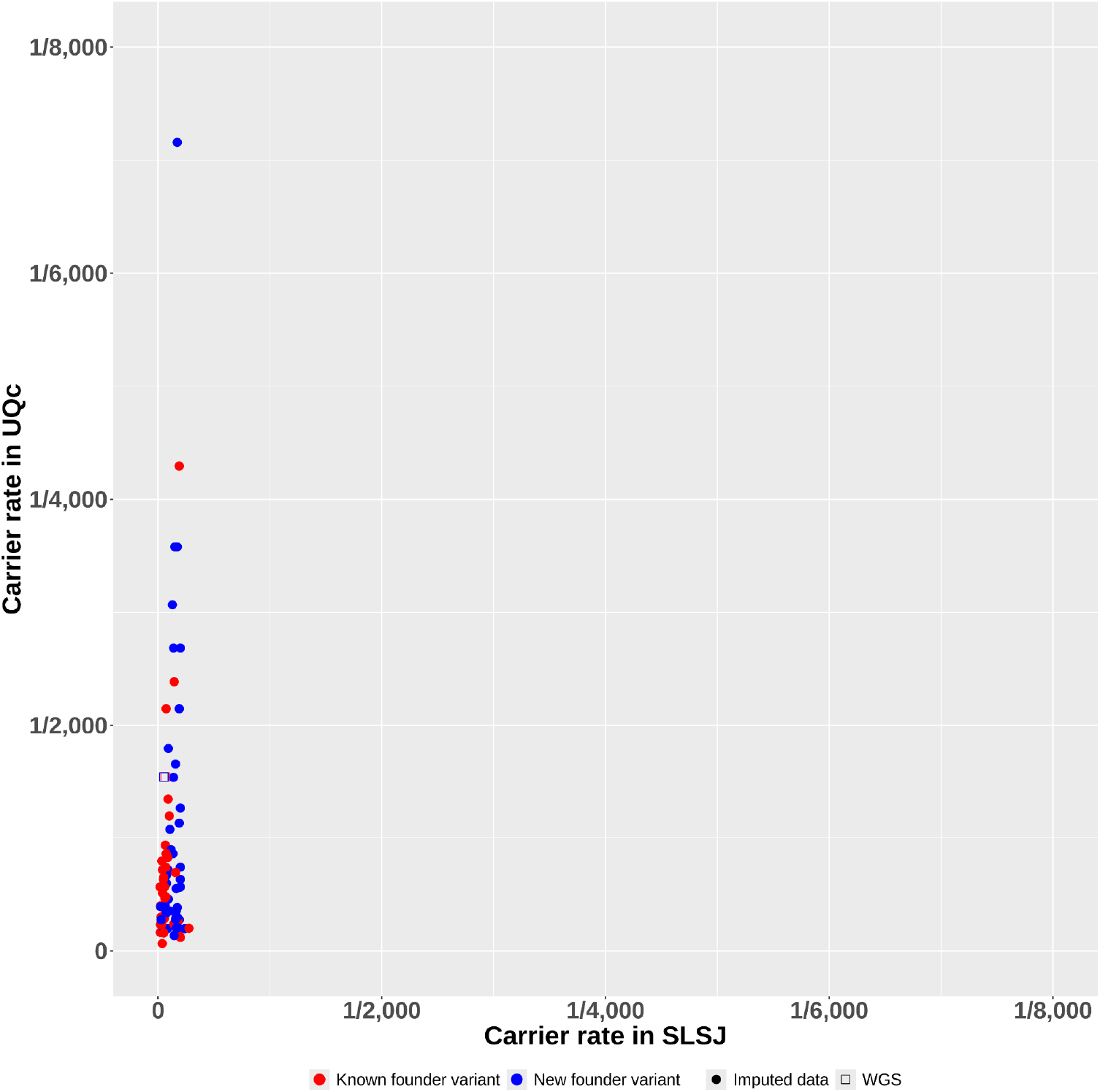
Carrier rates for founder variants. Only variants classified as founders in SLSJ or UQc or QcP are shown here (80 variants). When available, the CR from the imputed data was used; otherwise, the CR from WGS data was utilized.

**Fig. 4:**
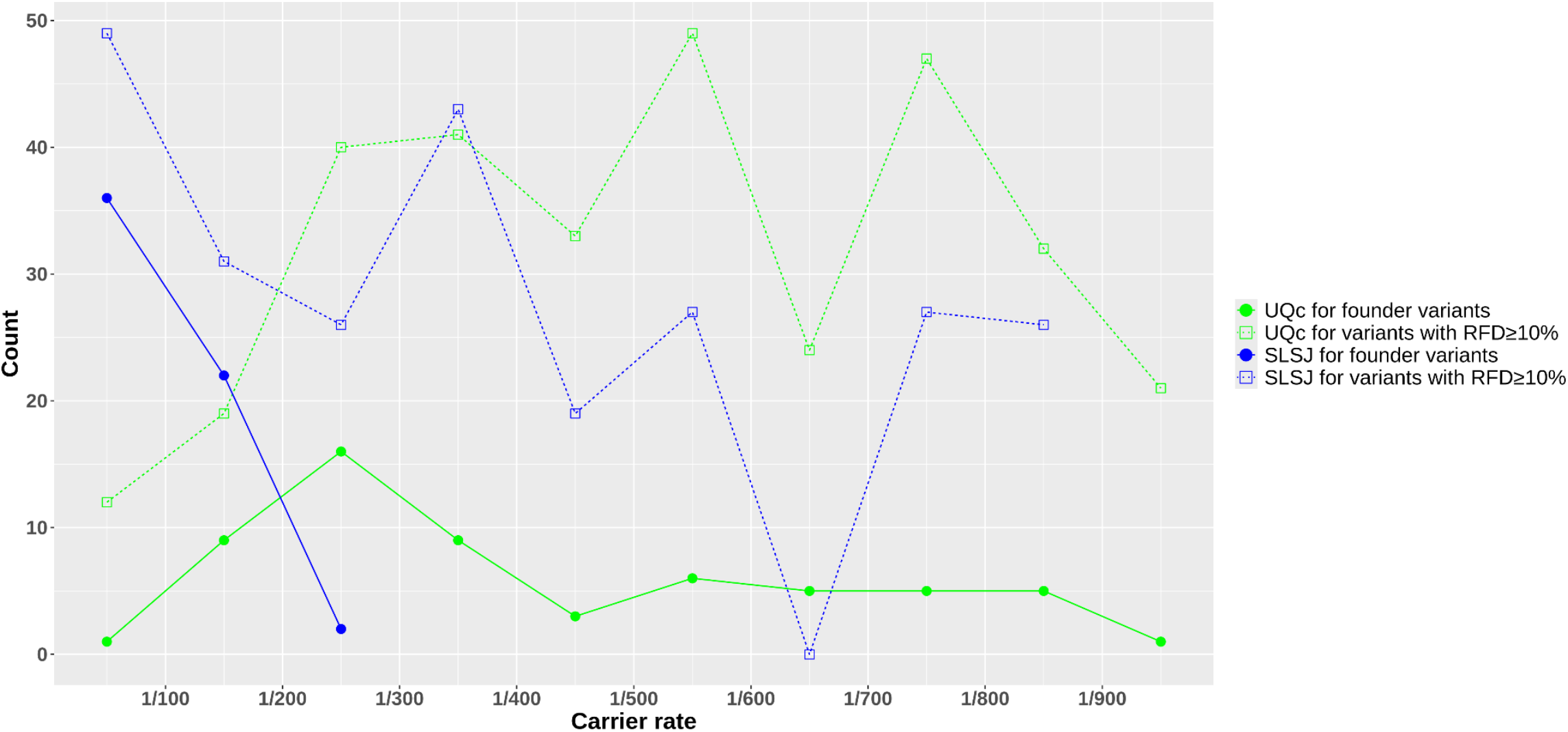
Number of variants in each carrier rate’s class. When available, the CR from the imputed data was used; otherwise, the CR from WGS data was utilized.

**Fig. 5:**
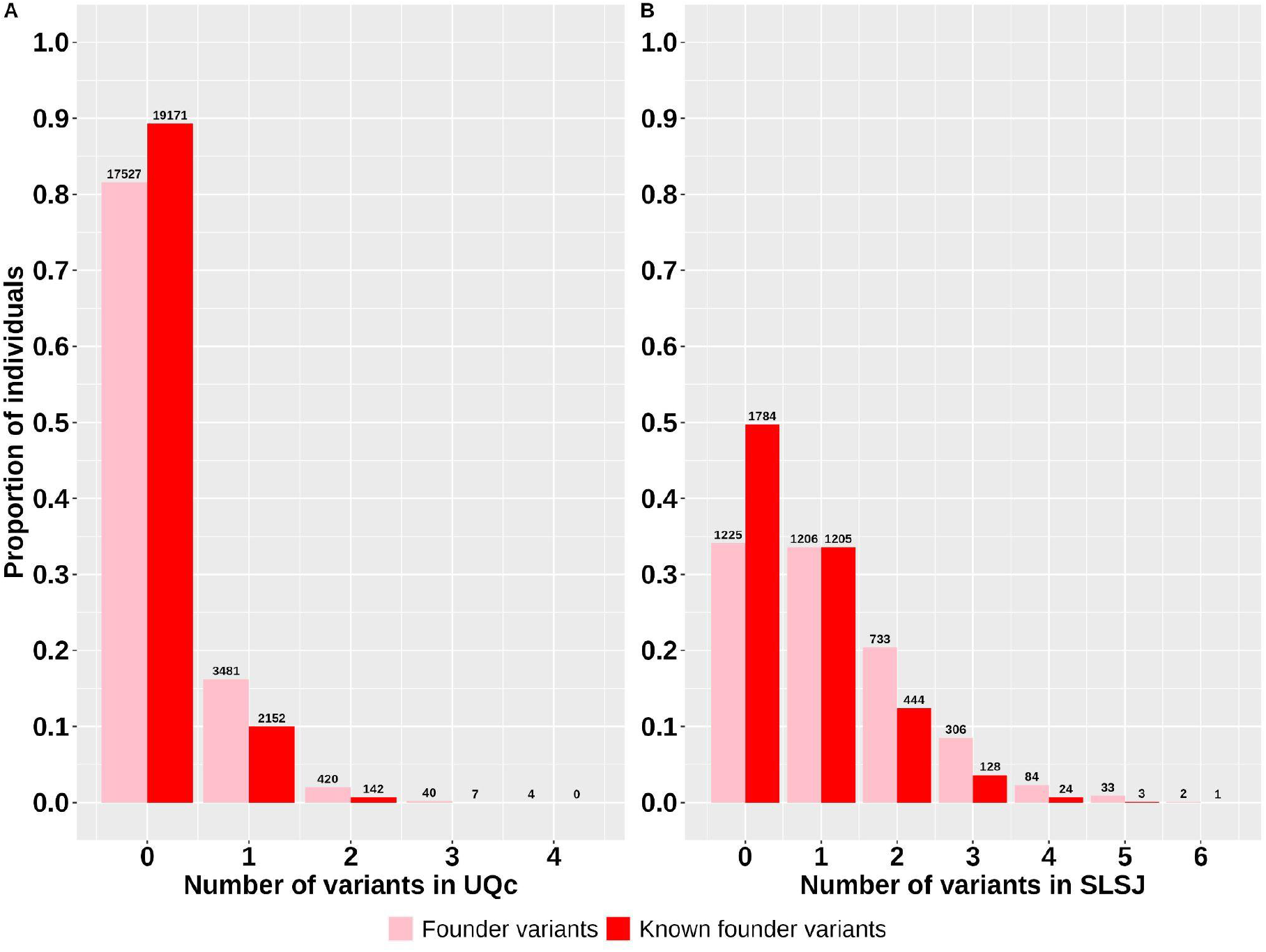
Proportion of individuals carrying founder variants in A) UQc and B) SLSJ. Only rare variants classified as founders in SLSJ or UQc or QcP are shown here (80 variants).

**Fig. 6:**
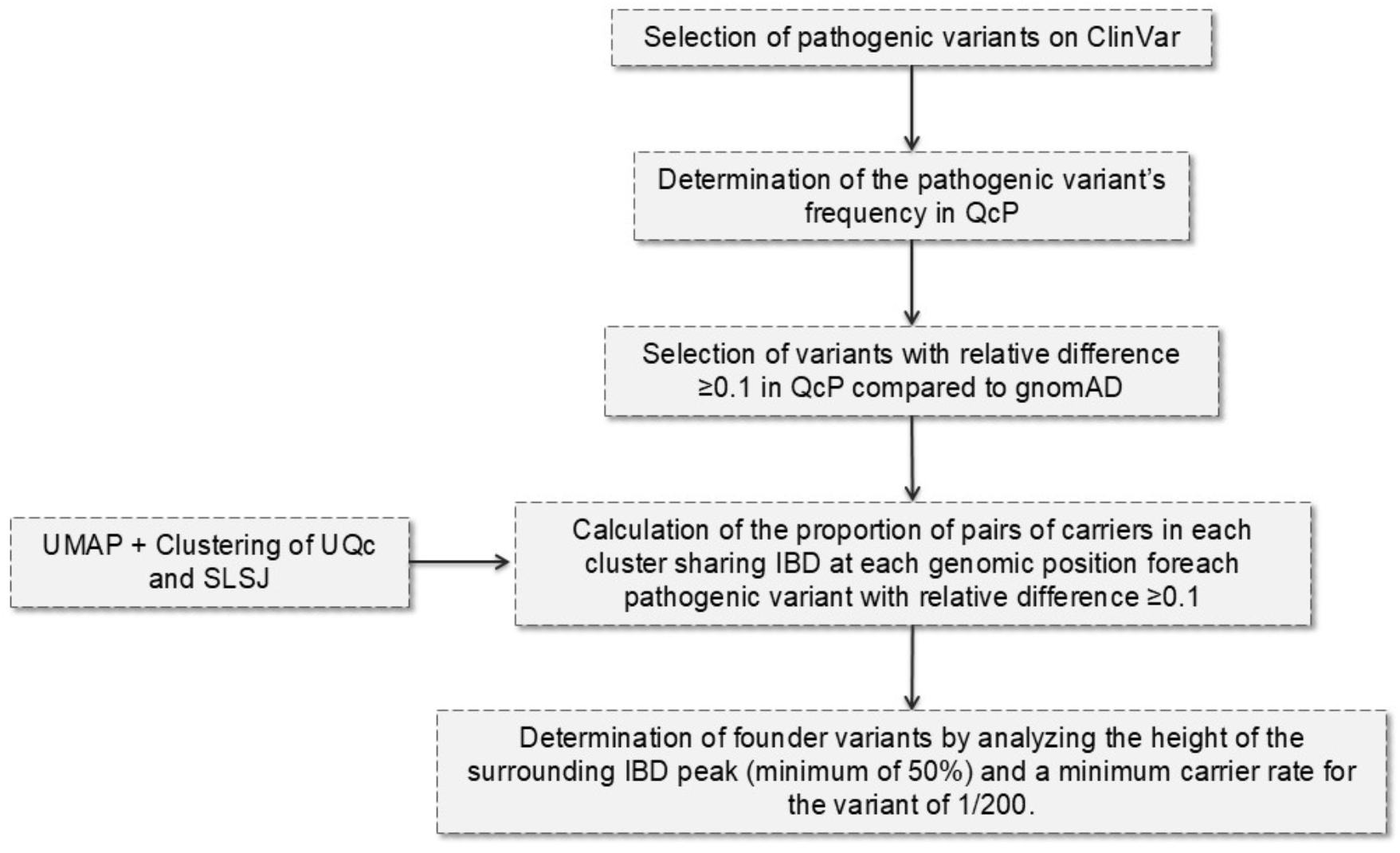
Simplified view of the flow of the analysis.

### Clinical validation

To confirm that our method identifies clinically relevant variants in the SLSJ population, we requested clinical experts to examine their databases seeking variants that segregate within families of patients presenting the corresponding phenotype. Table 3 presents the variants found in patients from the Medical Genetics service and the CMNM in addition to CARTaGENE phenotypes provided. Of note, 3 of the variants identified herein (Alstrom syndrome (550627), Multiple acyl-CoA dehydrogenase deficiency (459956) and Joubert syndrome 9 (217607)) would be good candidates to include in an ongoing effort for designing a new carrier test for the SLSJ population in the Medical Genetics service of the CIUSSS of the SLSJ.

**Table 3:**
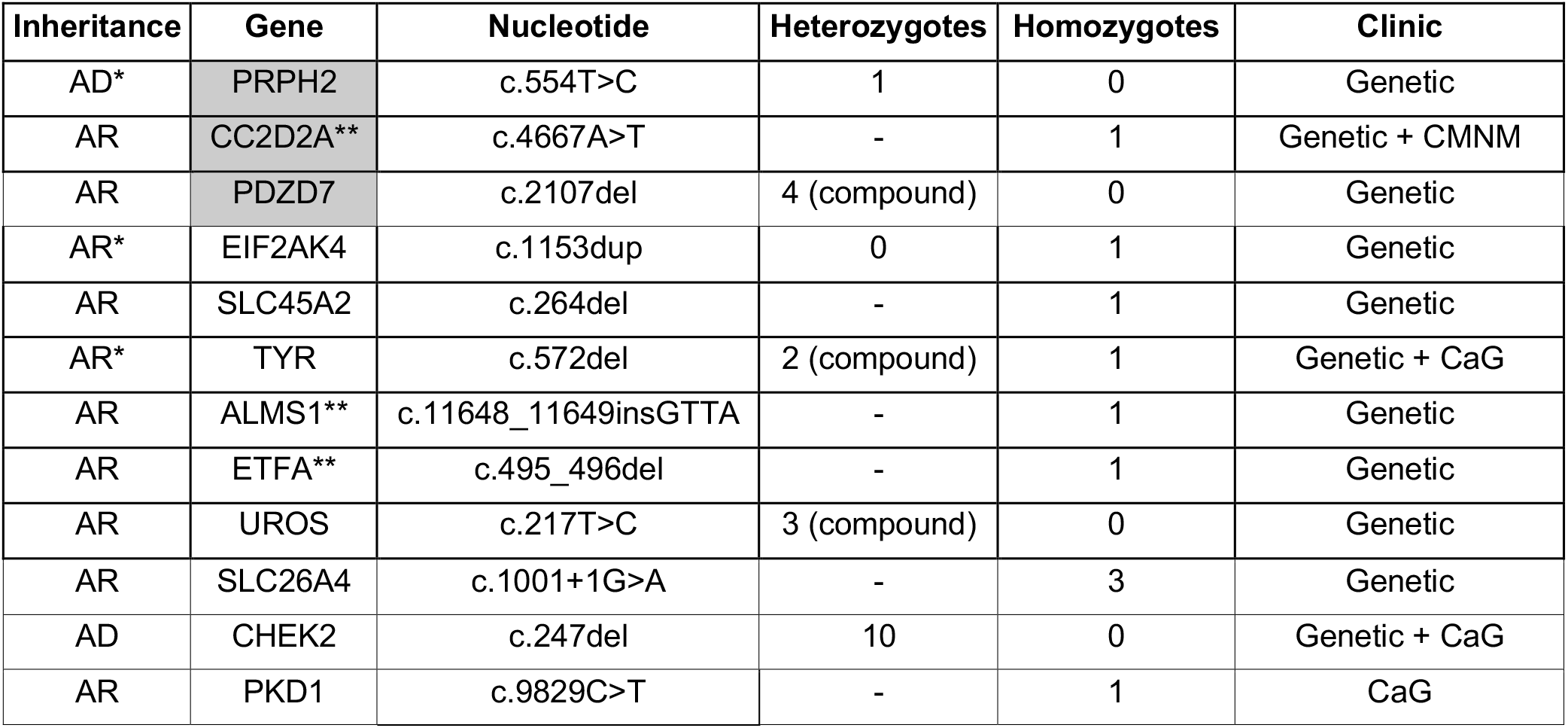
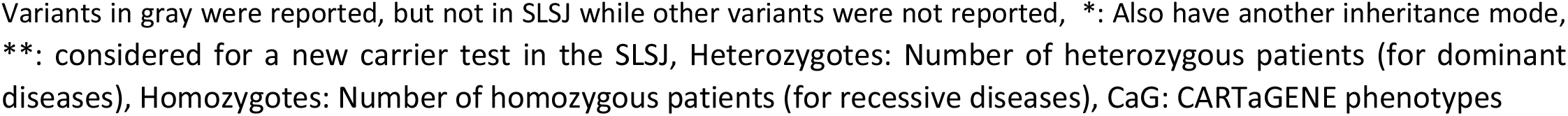
Clinical information for variants found in patients with corresponding phenotypes.

| Inheritance | Gene | Nucleotide | Heterozygotes | Homozygotes | Clinic |
| --- | --- | --- | --- | --- | --- |
| AD* | PRPH2 | c.554T>C | 1 | 0 | Genetic |
| AR | CC2D2A** | c.4667A>T | - | 1 | Genetic + CMNM |
| AR | PDZD7 | c.2107del | 4 (compound) | 0 | Genetic |
| AR* | EIF2AK4 | c.1153dup | 0 | 1 | Genetic |
| AR | SLC45A2 | c.264del | - | 1 | Genetic |
| AR* | TYR | c.572del | 2 (compound) | 1 | Genetic + CaG |
| AR | ALMS1** | c.11648_11649insGTTA | - | 1 | Genetic |
| AR | ETFA** | c.495_496del | - | 1 | Genetic |
| AR | UROS | c.217T>C | 3 (compound) | 0 | Genetic |
| AR | SLC26A4 | c.1001+1G>A | - | 3 | Genetic |
| AD | CHEK2 | c.247del | 10 | 0 | Genetic + CaG |
| AR | PKD1 | c.9829C>T | - | 1 | CaG |

Variants in gray were reported, but not in SLSJ while other variants were not reported, \*: Also have another inheritance mode, \*\*: considered for a new carrier test in the SLSJ, Heterozygotes: Number of heterozygous patients (for dominant diseases), Homozygotes: Number of homozygous patients (for recessive diseases), CaG: CARTaGENE phenotypes

## Discussion

In this study we aimed to identify potentially pathogenic variants found at higher frequency in the QcP and more specifically in the SLSJ region. Starting from 240,716 variants in ClinVar, we found 1,302 rare variants with RFD≥10% in Quebec compared to gnomAD non-Finnish Europeans (nfe). Among these 1,302 variants, we identified 80 that met our criteria to be classified as founders, with 38 being previously reported in the QcP. Note that we classified a founder variant as already known if it was reported at least once in the literature, either in the QcP or SLSJ or among French-Canadians. Consequently, we do not differentiate between the variants documented in studies for which the focus was on founder variants in Quebec^7–10^ and the ones identified in case reports over the years^13,15–27^.

In addition to taking the high carrier rate into account, we examined the shared IBD segments around the variant to identify variants associated with the founder effect. By doing so, we ensure that the variant originated from a single ancestor and was spread through drift in the population due to the founder effect. Additionally, keeping only variants with a CR of at least 1/200 avoids small familial or more recent sporadic increases in frequency that would not be attributable to a population founder effect. We also propose that variants with a high frequency in the population, but without a majority of pairs sharing a surrounding IBD segment, are likely the result of multiple introductions rather than a single one. Within the 42 variants previously characterized as founders in the literature and found in the present study^7–10^, 19 did not meet our criteria in any of the QcP, UQc or SLSJ groups primarily due to an insufficient number of carriers (Supplementary Table 1). Among these, 9 were documented alongside another founder variant for the same disease, leaving 10 variants without evidence of being real founder variants according to our criteria. Of note, all 15 variants associated with phenylketonuria identified in this study (4 of which were previously reported) had carrier rates below 1/200, which means that this condition was not classified as a founder disease in the present analysis. The same was observed for 5 other diseases (highlighted in gray in Table 1).

Regarding diseases caused by multiple variants, the literature often inaccurately reports that these are all at the same carrier rate in the population. It is rather the summed CR of all variants associated with a given disease which is reported. Indeed, by aggregating the carrier rates of the 10 variants present in our data for Cystic Fibrosis (Supplementary Table 1), we arrive at a final carrier rate of 1/16 which is very close to the one previously reported of 1/15^8^. However, each variant associated with one disease has its own CR and its own history. Among the 10 variants documented for Cystic Fibrosis^28^, our analysis reveals that 9 are nonfounders, including the 2 most frequent which seem to originate from multiple introductions in the population since less than half of pairs share IBD at the variant’s genomic location. The 7 other nonfounder variants are present with a carrier rate below 1/200. On the other hand, our findings suggest that 1 Cystic Fibrosis variant (CFTR c.489+1G>T) does meet our criteria. This variant is much more common in the SLSJ than elsewhere in Quebec indicating that it likely has risen in frequency during the Charlevoix-SLSJ regional founder effect. Thus, the increased prevalence of this disease in the SLSJ population can be partly attributed to multiple introductions of different genetic variants by various ancestors, with the Charlevoix-SLSJ founder effect being only one of several contributing factors.

In addition to confirming known founder variants, we also report for the first time 42 novel founder variants that, to our knowledge, have never been documented in the QcP. Some of these exhibit a high carrier rate, comparable to the most common known variants included in the carrier test offered to the population. These variants could potentially account for unreported rises in disease prevalence within the population, which suggests a potential underestimation of the overall prevalence of rare diseases in the SLSJ region as also reported in other populations with founder effects^29^. Indeed, adding the newly identified variants raises the proportion of individuals carrying at least one founder variant of 1.3 and 1.7 times in the SLSJ and in the UQc respectively. Establishing carrier rates plays a critical role in advancing precision medicine among populations with a founder effect^29^. In addition, it is a great proof-of-concept for larger initiatives to come in the field of precision medicine in regard to carrier frequency panels in larger populations.

Considering that we demonstrate an underestimation of the number of pathogenic variant carriers in SLSJ, which has been the focus of numerous studies on rare genetic diseases linked to the founder effect, we hypothesize that this phenomenon might also be present in other populations worldwide. Indeed, a rise in deleterious allele frequencies following range expansions has also been observed in other non-African populations^30^. Consequently, the number of individuals affected with a rare disease might be underestimated in many countries or local communities. Our population cohort’s approach could be applied in other worldwide populations at low costs thus helping in enhancing and fastening the molecular diagnosis of patients.

When comparing pathogenic variant frequencies between gnomAD and the QcP, we observed almost 4 times more deleterious variants with an RFD ≥10% in the latter (Supplementary Fig.2) in line with the higher deleterious mutation load previously observed in rapidly expanding populations^31^. The present study also demonstrates the higher pathogenic mutational load of individuals from the SLSJ region compared to UQc not only for founder variants, but also for variants with an RFD ≥10%. It seems that the UQc group has a higher number of unique variants with an RFD ≥10%, while the SLSJ individuals are more likely to carry two or more variants. Indeed, 42% of variants with an RFD ≥10% in the QcP were lost in the SLSJ, although some of them might be too rare to be observed in the SLSJ due to the smaller sample size. As for the 80 founder variants, again a greater mutational load in addition to a higher number of variants with CR above 1/200 are observed in the individuals from the SLSJ compared with those from the UQc. It can be concluded that the higher mutation load in the SLSJ individuals is mainly caused by an overrepresentation of variants with a CR greater than 1/200. This is the result of the genetic bottleneck of the SLSJ followed by a very rapid population expansion, 5 times greater than the one observed in the whole Quebec for the same period^32^, which represents one of the strongest regional founder effects in Quebec^6^. Some founders in this region contributed a lot to the present population^33^ and therefore could have introduced an allele in the population that would reach such a high frequency^34^. Moreover, it was demonstrated that the first SLSJ settlers had an increased fitness^35^ which could have contributed to increasing deleterious allele frequencies^36,37^ possibly due to increased drift and relaxed selection^38^.

To validate that the variants identified in this study are associated with specific diseases, we searched clinical databases and CARTaGENE phenotypes for patients carrying those variants who have been diagnosed with the corresponding disease. Notably, 12 of the variants identified in this study were detected in patients from the CIUSSS of SLSJ clinics and/or in CARTaGENE. Those variants have not been previously reported in the SLSJ, although 3 of them (PRPH2 c.554T>C, CC2D2A c.4667A>T, PDZ7 c.2107del) were reported in the French-Canadian population.

This study has certain limitations. Firstly, the sample size of the WGS data may be insufficient to accurately estimate the frequency of variants in the population. Therefore, we chose to work with imputed data, which includes a significantly larger number of individuals. To achieve the most accurate representation of our data, especially given our focus on rare variants, we performed imputation using our WGS data rather than a global worldwide reference panel. However, we acknowledge that imputed data may not be as reliable as WGS or genotyping. Therefore, we compared the WGS data with the imputed data for the same individuals and excluded any imputed variants that were false positives or not present in the WGS data. This sometimes could affect comparisons of CR and cumulative CR for different variants due to differences in sample sizes between both data types. We also excluded any individual whose cluster in WGS did not match the one in the imputed data clustering based on the UMAP. Also, our definition of a founder variant is stringent with a CR of at least 1/200, especially for the SLSJ region where the sample size is smaller and this CR could not be reached for the WGS. As a result, some less common but genuine founder variants might be missed. Lastly, our conclusions regarding the mutation load apply only for the 1,302 variants with an RFD ≥10% in QcP as we did not assess the load on all variants with a confirmed pathogenicity in ClinVar.

In conclusion, this study demonstrates a greater mutation load for founder variants and for variants with an RFD ≥10% in the SLSJ region compared to urban Quebec areas due to its stronger founder effect. In fact, this load is driven by more numerous variants present at a higher frequency in the SLSJ population. In addition to confirming previously described pathogenic variants using a population cohort instead of a clinical approach, we found variants associated with diseases that were not yet described in Quebec or in the SLSJ. These findings might be crucial for clinicians to shorten the patients’ diagnostic odyssey and reduce the economic burden associated with undiagnosed rare diseases. This could help improve the management of patients and, for some or them, enhance their quality of life and slow disease progression as appropriate treatments could be offered earlier. With this information, precision medicine can implement targeted genetic screening programs, allowing for early detection of inherited conditions that are more prevalent due to the founder effect. This enables tailored prevention strategies, personalized treatments, and risk-reduction measures that are specific to the genetics of the population. Additionally, pharmacogenomics can benefit from this knowledge by optimizing drug therapies based on genetic susceptibilities. Ultimately, understanding carrier rates in populations with a founder effect helps healthcare providers offer more precise and effective medical care, enhancing outcomes for both individuals and the community. Indeed we identified 30 patients carrying 12 causal variants that have not been previously reported in SLSJ. The underestimation of pathogenic mutational load might also happen in other populations as a result of range expansions and rare diseases might be much less rare than anticipated. In an era of precision medicine with at least 10% of the population affected by rare diseases, it is crucial to adopt new approaches to enhance and fasten the molecular diagnosis of rare diseases.

## Data and Methods

This study was approved by the University of Quebec in Chicoutimi (UQAC) ethics board. The approval for the secondary use of anonymized samples coming from the provincial screening testing was obtained from the CIUSSS of the SLSJ Direction of professional services. Written informed consent for the use of saliva samples were obtained from participants.

### Cohort

The CARTaGENE cohort^39^ (https://cartagene.qc.ca/) used in this study includes WGS of 2,184 and genotyping of 29,337 participants. Individuals aged between 40 and 69, residing in 6 distinct cities (Montreal, Quebec City, Trois-Rivières, Sherbrooke, Gatineau, Saguenay), were recruited between 2009 and 2015, regardless of their birthplace. The CARTaGENE cohort also includes a wide range of phenotypes, among which are the occurrence of a disease. The genotype and WGS data quality control are described here (https://cartagene.qc.ca/files/documents/other/Info_GeneticData3juillet2023.pdf). All genomic data were alignmed on the GRCh38 genome assembly.

### Genotypes cleaning and imputation

To increase our sample size and achieve more accurate carrier rates, we imputed the 6 different CARTaGENE genotyping chips using SHAPEIT5^40^ and IMPUTE5^41^. The individuals were genotyped on different arrays (Omni 2.5, GSAv1 + Multi disease panel, GSAv1, GSAv2 + Multi disease panel, GSAv3 + Multi disease panel, GSAv2 + Multi disease panel + addon and Affymetrix Axiom 2.0) (https://cartagene.qc.ca/files/documents/other/Info_GeneticData3juillet2023.pdf), and were cleaned and merged as follows. Each dataset was cleaned separately using PLINK software v1.9^42^, ensuring individuals with at least 95% genotypes among all SNPs were retained. At the SNP level, we retained SNPs with at least 95% genotypes among all individuals, located on the autosomes and in Hardy–Weinberg equilibrium p > 10^−6^ (calculated on each dataset).

The imputation was performed on each genotyping batch separately using the 2,184 CARTaGENE WGS as reference to enhance our capacity to identify rare variants within our population. All batches were then merged and the final imputed dataset includes 29,353 individuals. A postimputation quality control filter was applied on each individual imputed batch to remove variants with an imputation quality score <0.3 for the PCA and UMAP.

### UMAP and clustering according to individuals’ origin

As a reminder, the CARTaGENE individuals’ recruitment site was based on their current residence rather than their birth place, even though many individuals live in their place of birth. For the purpose of this study, we needed to identify the most related individuals based on genetics, regardless of where they were recruited. To do so, a PCA was performed using PLINK on the WGS SNPs with a minor allele frequency (MAF) of at least 5% and after removal of SNPs with more than 2% missing individuals and in LD. We retained only biallelic SNPs within the accessibility mask^43^, resulting in a total of 90,073 remaining SNPs. We also filtered out individuals with more than 2% missing SNPs resulting in 2,166 individuals remaining. A UMAP^44^ was then performed on the 3 first PCs (determined by the scree test) with the R umap library v0.9.2.0 (Supplementary Fig.3). This technique was proven efficient to reveal fine-scale population structure^45^. K-means clustering was then employed to create 3 clusters, aiming to retain as many individuals from the SLSJ as possible, given its limited sample size, especially for the WGS data. We also intended to choose individuals with the strongest ancestry connection to the region. Based on the recruitment place (Supplementary Fig.3A), we could see that the majority of the CARTaGENE participants recruited from the SLSJ region belongs to the red cluster (Supplementary Fig.3B). In fact, the red cluster of the WGS data encompasses 90% of the individuals who were recruited from the SLSJ region. We identified 314 individuals originating from the SLSJ region (red cluster) who were recruited in different places and 1,538 individuals from the other urban Quebec regions (UQc) (green cluster), for a total of 1,852 for the QcP (green and red clusters). Clusters were also defined on imputed data as described above on pruned SNPs at 5% frequency or more keeping 5 PCs for the UMAP, leaving 3,589 individuals in the SLSJ (red cluster) and 21,472 in the UQc (green cluster), for a total of 25,061 in the QcP (Supplementary Fig.4). For the imputed data, the red cluster encompasses 84% of the individuals who were recruited from the SLSJ region. We ensured consistency of individuals in clusters between the WGS and imputed data by removing 27 samples that exhibited mismatches, likely due to sample mix-ups in one dataset or because they were at the boundaries of both clusters. This method ensures that individuals have a common genetic background and was shown to be helpful in uncovering rare variants with smaller sample sizes^46,47^.

### Selection of pathogenic variants with relative frequency difference ≥ 10%

Variants’ classification was extracted from the ClinVar database version of June 24, 2024^48^. Only variants classified as: Pathogenic, Likely pathogenic, and conflicting (both pathogenic and likely pathogenic variants), as well as SNPs, insertions and deletions (indels), were included in the analysis whereas repeat expansions were excluded. Furthermore, variants with the following review status were removed: no assertion criteria provided, no classification provided, no classification for the individual variant. Additionally, we incorporated all variants referenced as founder variants in previous studies^7–10^, regardless of their status on ClinVar. Therefore, we obtain a list of 240,716 variants.

We calculated the variants’ frequency in the WGS and imputed data using PLINK v1.9 for the individuals originating from the SLSJ, UQc and QcP (both clusters) inferred by the clustering. Whenever a variant was present and was not a false positive in the imputed data, we used the imputed variant frequency, otherwise, we relied on the WGS variant frequency. Only WGS variants with less than 10% missing individuals were kept. Notably, the frequencies of variants show strong correlation between both data types (Supplementary Fig.5). The gnomAD frequencies for the non-Finnish Europeans (non_topmed_nfe) were directly extracted from gnomAD genomes v3.1.2^49^. To calculate the relative frequency difference (RFD) of a variant in the QcP compared to gnomAD nfe, we used the following formula:

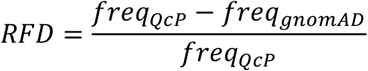

Knowing that:

- *freq*_*Qcp*_ corresponds to the frequency of the variant in the *Qcp* population.
- *freq*_*gnomAD*_ corresponds to the frequency of the variant in the gnomAD non-Finnish Europeans.

We fixed a minimum RFD threshold of 0.1 to make sure it encompasses all variants that could be of interest. We detected 1,304 potentially pathogenic variants in the WGS of 1,852 individuals within the QcP that reached RFD≥10% compared to gnomAD nfe. Since we are focussing on rare variants, we removed 2 variants with a MAF ≥ 5% leaving 1,302 rare variants with RFD≥10% in the QcP.

### Estimation of carrier rate

We directly counted the number of heterozygotes for each variant and determined the CR by calculating the inverse of the frequency of the heterozygous individuals as follows:

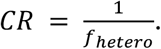

### Selection of founder variants

Furthermore, we established criteria for a variant to be called as founder. The number of individuals carrying the variant must be adequate to avoid false signals or misinterpretations while also being high enough to be clinically relevant; thus, the target CR was set to 1/200. Hence, we set the minimum threshold at 5 (1/63), 8 (1/192) and 10 (1/185) individuals carrying the variant for WGS of SLSJ, UQc and QcP respectively. However, for imputed data, we set the threshold to reach a CR of 1/200 which represents 18, 108 and 126 individuals for the SLSJ, UQc and QcP. Furthermore, to be called as founder, a variant must show a proportion of pairs of carriers sharing an IBD segment around the variant (see next section) of at least 0.5.

### IBD segments inference and sharing

All cleaned genotyping batches (excluding the Affymetrix chip due to its poor SNPs intersection with other Illumina chips) were combined and only the intersecting common SNPs were kept. After the merge, individuals with less than 95% genotypes among all SNPs and SNPs with less than 95% genotypes across all individuals were once again filtered out. The final dataset comprises 148,200 SNPs and 28,358 individuals.

We then inferred IBD segments on phased genotypes using refinedIBD^50^ version 17Jan20 and Beagle version 18May20. Subsequently, the segments were merged using the merge-ibd-segments 17Jan20.102 tool. We retained only IBD segments of 2Mb or longer and with a LOD score greater than 3.

We then examined the genome-wide IBD segments shared among pairs of individuals carrying a specific pathogenic variant with RFD≥10% and determined the proportion of pairs sharing IBD at each genomic position. We considered the variant as a founder variant if this proportion reached a minimum of 0.5, indicating that half of the pairs share IBD at the variant’s location. Considering that a founder variant usually originates from a single ancestor in a population with a founder effect^34^, and within a relatively recent time frame (with the first permanent settlement in Quebec starting in 1608), this variant is often inherited with other variants in LD. Therefore, examining the IBD sharing around a variant is a dependable method to confirm its status as a true founder variant.

### Calculation of genome-wide relatedness

To validate that our findings are not subject to potential high relatedness biases, we conducted genetic kinship calculations among individuals carrying the same variant. To do so, we assessed the total length of IBD segments shared between pairs of individuals divided by twice the length of the genome (to account for the diploid human genome). If the resulting percentage exceeded 50%, it suggested a potential first-degree relationship between the individuals. Detecting these relationships could reveal potential biases in our IBD analysis, particularly if individuals shared a recent common ancestor. In such cases, the IBD segments they share may not accurately represent a founder effect, but instead, direct familial transmission from the recent ancestor. For carriers of founder variants, the average whole genome IBD sharing between pairs of individuals with the same variant never exceeds 5%. This suggests that the observed proportion of individuals sharing IBD around the founder variants is not attributable to close relatedness.

### Clinical data

The patient group consisted of individuals residing in SLSJ during the assessment, all of whom had genetic disorders. They were clinically evaluated at the Medical Genetics service and the CMNM of CIUSSS of SLSJ. Their DNA samples were analyzed in certified clinical molecular laboratories as part of the clinical testing and genetic evaluation process. A review of internal databases and the patients’ medical records enabled the identification of patients who were homozygous, compound heterozygous, or heterozygous for autosomal recessive or dominant variants.

### Experimental validation of carrier rates

To validate the estimated carrier rates derived from the WGS and imputed data, we randomly selected 1,000 individuals from the SLSJ who had consented to the storage of their anonymized DNA samples for research purposes. We chose 2 founder variants, DOK7 c.1124_1127dup and CTNS c.414G>A, for genotyping due to their high CR observed in the SLSJ in the present study (1/21 and 1/63, respectively). They were genotyped using custom TaqMan genotyping assays (catalog #4332072; Applied Biosystem Inc). We designed the assays with specific probes targeting each allele using the Primer Express software. We extracted the DNA from buccal swab samples using DNA extract all kit (catalog #4402616; Applied Biosystem Inc) following the manufacturer recommendations. In brief, 22.0 μl of Lysis solution was added to 7.0 μl of buccal swab emulsions, then incubated for 3 minutes at 95°C in a thermocycler. 22.0 μl of DNA stabilizing solution was then added to the mix. For amplification and detection, the manufacturer recommendations were followed. In brief, for each reaction 1.75 μl of sterile water, 3.10 μl of GTXpress Master Mix (catalog #4401892; Applied Biosystem Inc), 0.15 μl of TaqMan assay, and 1.2 μl of DNA. Analysis was carried out in a 96-well plate and samples were amplified on a 7500 Fast Real-Time PCR thermocycler (Applied Biosystems Inc). The amplification conditions were as follows: 1: 60°C for 1 min with fluorescence acquisition, 2: 95°C for 20 s, 3: 95°C for 3 s, and 60°C for 30 s with fluorescence acquisition (step 3 was repeated 40 times), 4: 60°C for 1 min with fluorescence acquisition. The genotypes were called using 7500 Software v2.0.1 (Applied Biosystem Inc) after visual inspection of the amplification data. For the first variant, 33 samples were excluded from the analysis due to unsuccessful PCR amplification (N=967), while for the second variant, 10 samples were excluded from the analysis (N=990).

## Abbreviations

SLSJ: (Saguenay–Lac-Saint-Jean)
UQc: (Urban Quebec regions)
QcP: (Quebec province)
WGS: (Whole-genome sequencing)
IBD: (Identical-by-descent)
CR: (Carrier rate)
MAF: (Minor allele frequency)
RFD≥10%: (Relative frequency difference of at least 10%)
LD: (Linkage disequilibrium)
CaG: (CARTaGENE cohort)

## Data availability

Quebec genotype, imputed and WGS data are available via an independent data access committee by the CARTaGENE cohort (https://cartagene.qc.ca/en/researchers/access-request.html).

## Code availability

The code used for this study can be found in the following GitHub repository: https://github.com/ElisaMichel/Founder_variant_2024.

## Acknowledgements

Funding for SLG was provided by the Canada Research Chair in Genetics and Genealogy. LB and CG were funded by the Research Chair in *Génétique et parcours de vie en santé*.

