## Supplementary file for "Rare diseases load through the study of a regional population"

***Supplementary Table 1: Variants with RFD≥10%.***

***Supplementary Table 2: Variants previously described in the population not found here.***

***Supplementary Table 3: Experimental reassessment of CR***

| Gene | Nucleotide | This study CR | Reassessed CR | Lower CI | Higher CI |
|---|---|---|---|---|---|
| DOK7 | c.1124_1127dup | 1/21 | 1/29 | 1/39 | 1/21 |
| CTNS | c.414G>A | 1/63 | 1/43 | 1/68 | 1/26 |

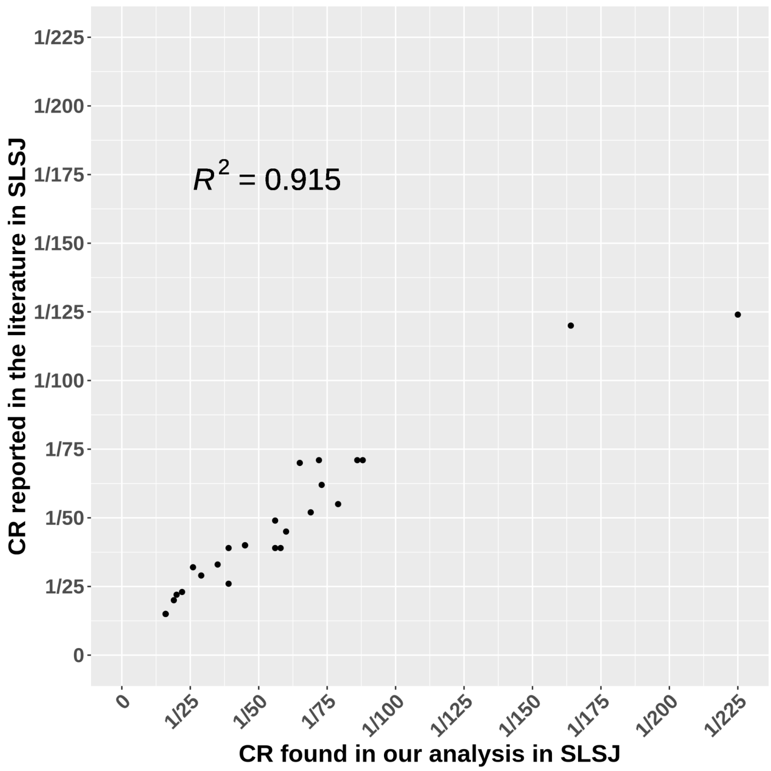


***Supplementary Fig.1: Comparison of the variants' carrier rates reported in SLSJ and found in our analysis.*** *When available, the aggregated CR was used; also, if available, the CR from the imputed data was used; otherwise, the CR from WGS data was utilized.*

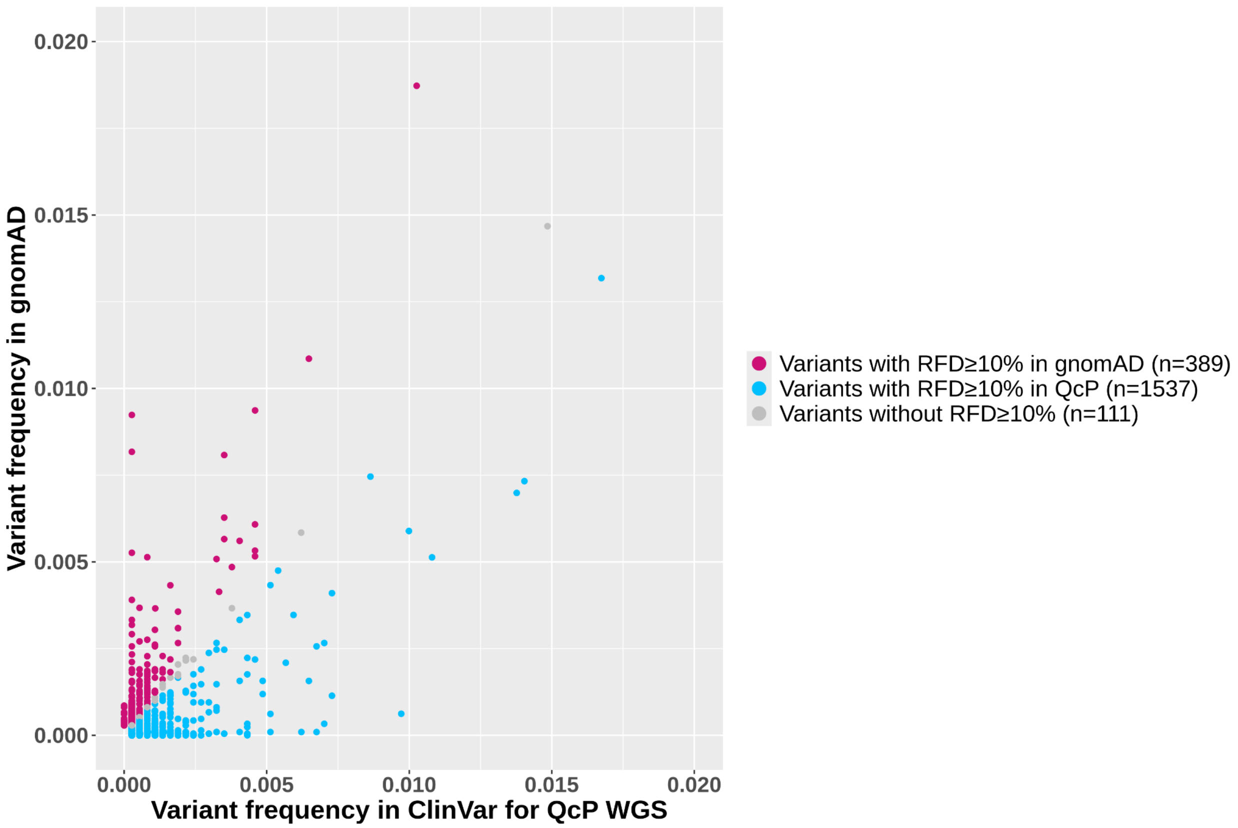


***Supplementary Fig.2: Comparison of the variant frequencies in gnomAD and WGS QcP.*** *ClinVar pathogenic variants with MAF < 2% present in both gnomAD and QcP WGS were included in this graph.*

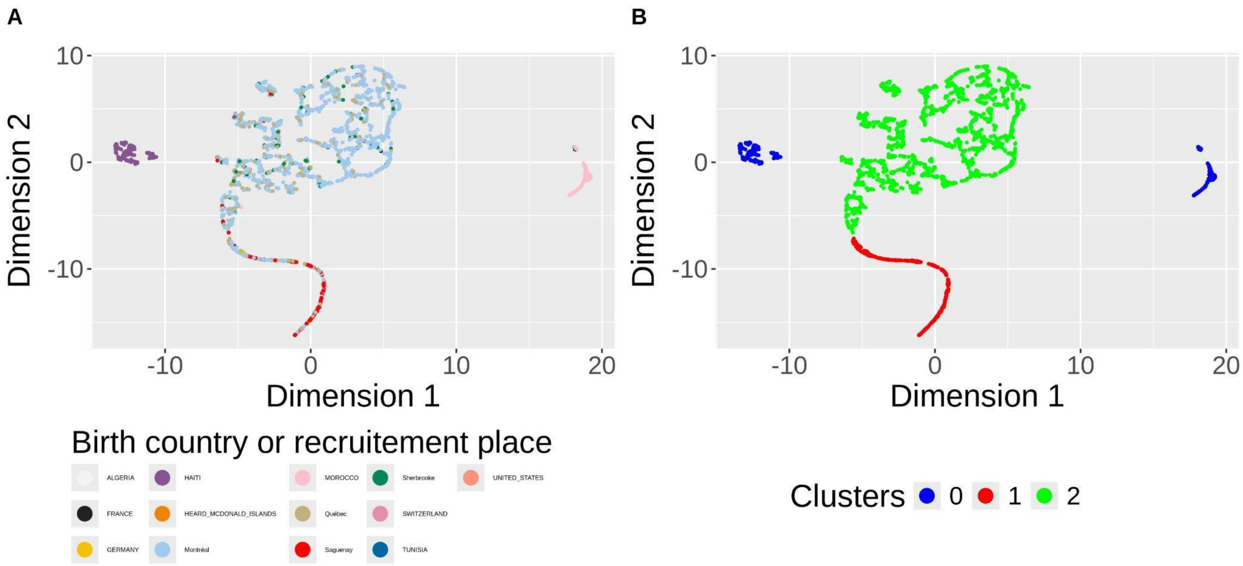


***Supplementary Fig.3: UMAP of WGS data.*** *UMAP are coloured according to A) the recruitment region or country of birth and B) the k-means clustering.*

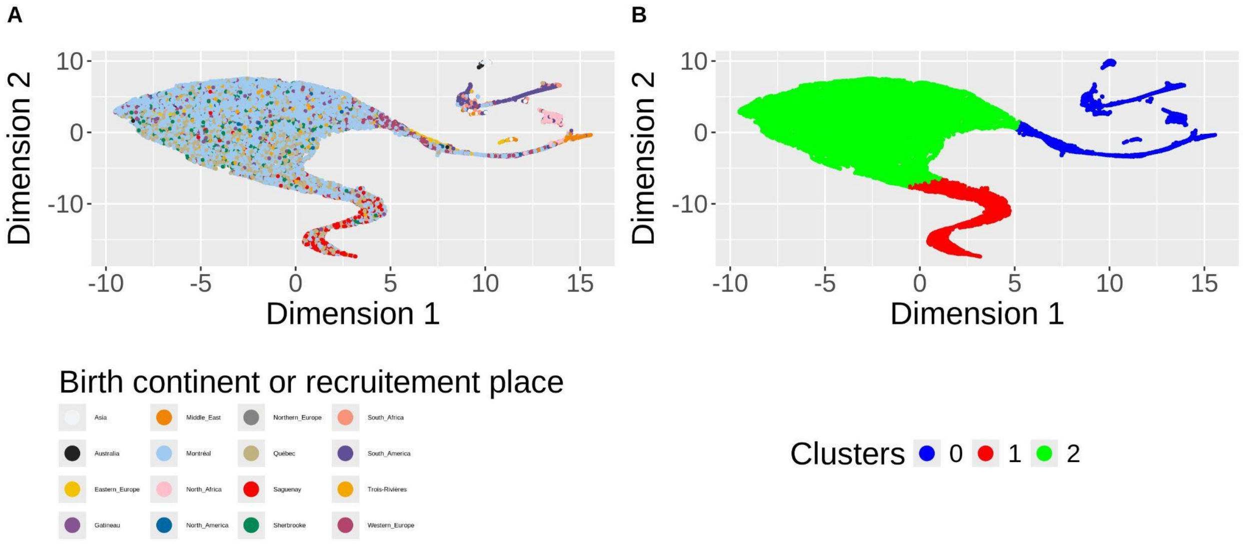


***Supplementary Fig.4: UMAP of imputed data.*** *UMAP are coloured according to A) the recruitment region or continent of birth and B) the k-means clustering.*

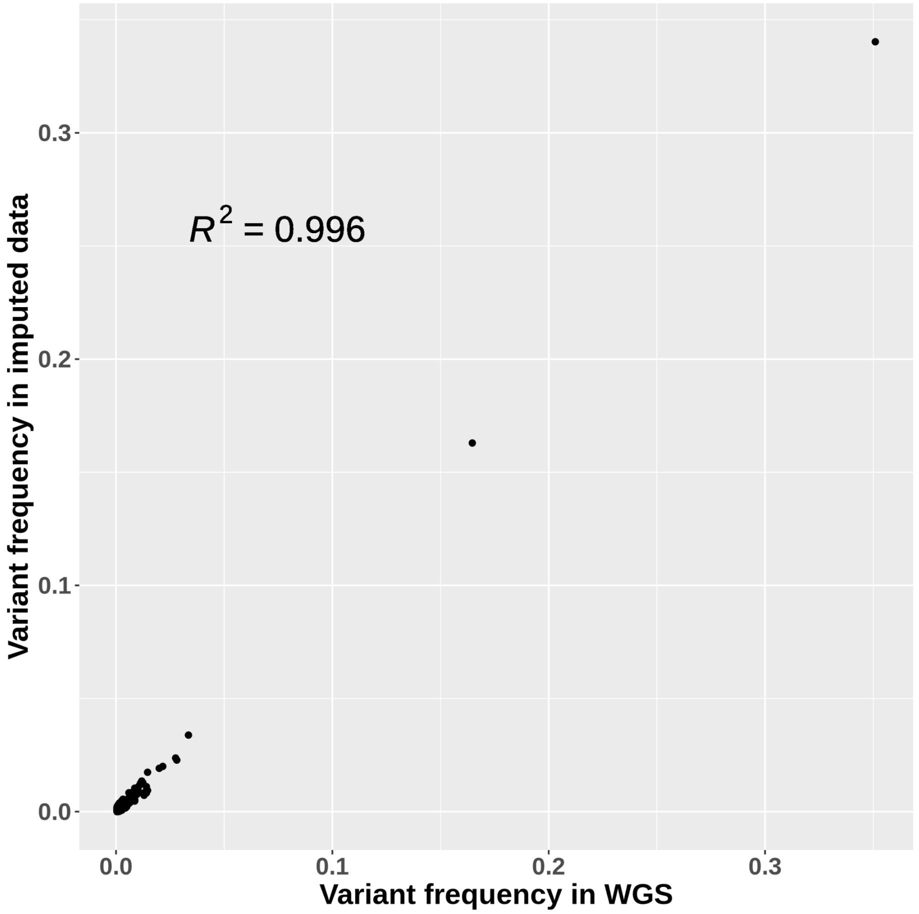


***Supplementary Fig.5: Correlation between imputed and WGS variants' frequency in QcP.***
